# Longitudinal clinical, physiological and molecular profiling of female metastatic cancer patients: protocol and feasibility of a multicenter high-definition oncology study

**DOI:** 10.1101/2025.05.14.25327544

**Authors:** Leonardo D. Garma, Sonia Pernas, David Vicente Baz, Rosario García Campelo, Josefa Terrasa, Berta Nasarre, Ruth Vera García, Begoña Bermejo, Santiago González Santiago, Bartomeu Fullana, Mario Rodríguez, Manuel Fernandez Bruno, Cristina Reboredo Rendo, Antonia Perelló, Desirée Jiménez, Juan Antonio Guerra, Virginia Arrazubi, Susana de la Cruz, Juan Miguel Cejalvo, Cristina Tebar, Elena Núñez Berrueco, Berta Hernández-Marín, Jacobo Rogado, Pablo M Olmos, Leire P. Arbaizar, Miguel A. Villalba Oliva, Yolanda Madrid, Jon Sanz-Landaluze, Gustavo Moreno-Martín, Aída Morillas, Antonio Lopez-Alonso, Alfonso Rodríguez-Patón, Ramón Colomer, Antonio Artés, Miguel Quintela-Fandino

**Affiliations:** Breast Cancer Clinical Research Unit, Centro Nacional de Investigaciones Oncologicas (CNIO), Madrid, Spain; Institut Catala d’Oncologia (ICO)-Institut d’Investigació Biomèdica de Bellvitge, Barcelona, Spain; Hospital Virgen de la Macarena, Sevilla, Spain; Medical Oncology Service, University Hospital A Coruña (XXIAC-SERGAS), A Coruña, Spain; Department of Medical Oncology, Hospital Son Espases, Palma de Mallorca, Spain; Hospital Universitario de Fuenlabrada, Fuenlabrada, Spain; Department of Oncology, University Hospital of Navarra, Instituto de Investigación Sanitaria de Navarra, IdISNA, Navarra, Spain.; Medical oncology, Hospìtal Clínico Universitario de Valencia, Biomedical Research Institute INCLIVA, Medicine Department, Universidad de Valencia, Valencia, Spain; Hospital San Pedro de Alcántara, Cáceres, Spain; Departamento de Inteligencia Artificial. Universidad Politécnica de Madrid (UPM), Madrid, Spain; Medical Oncology Division, Hospital Universitario La Princesa, Madrid, Spain; Dept Signal Theory & Communications, Universidad Carlos III de Madrid, Madrid, Spain; Universidad Complutense de Madrid (ROR 02p0gd045). Facultad de Ciencias Químicas. Ciudad Universitaria, 28040 Madrid, España; Department of Medicine, Universidad Autónoma de Madrid (UAM), Madrid, Spain; Chair of Personalised Precision Medicine, Universidad Autonoma de Madrid (UAM – Fundación Instituto Roche), Madrid, Spain; Evidence-Based Behavior, Madrid, Spain; Instituto de Investigación Sanitaria Gregorio Marañón, Madrid, Spain; CIBERSAM, Spain, Barcelona, Spain

## Abstract

**PURPOSE:** A substantial proportion of patients receiving genomically matched therapies do not achieve clinical benefit, underscoring the influence of non-genetic factors such as microbiome composition, metabolic state, physiological parameters, and lifestyle behaviors on cancer outcomes. High-Definition Oncology (HDO) proposes integrating longitudinal, multi-modal patient data—spanning clinical, molecular, physiological, and behavioral domains—to enable truly individualized cancer care. The present manuscript describes the design, methodological framework, and initial feasibility results of the HDO study, which aims to evaluate the feasibility of harmonized, deep longitudinal profiling across clinical, molecular, physiological, and behavioral domains in women with metastatic cancer.

**PATIENTS AND METHODS:** We initiated a prospective, multicenter observational study (HDO study; NCT06590506) enrolling 300 female patients with newly diagnosed metastatic breast, lung, or colorectal cancer. Here, we report the study design, standardized workflows, prespecified feasibility criteria, and early internal pilot results. Eleven data modalities are collected longitudinally, including tumor and germline genomics, germline epigenomics, gut microbiome, blood and stool metabolomics and proteomics, exposome characterization, wearable-derived physiological monitoring, digital footprint assessment, medical imaging, and patient-reported outcomes. Standardized workflows govern clinical procedures, data acquisition, biospecimen processing, and quality control across all participating sites.

**RESULTS:** Feasibility was evaluated in the first 30 participants (10% of planned accrual). Patients completed 100% of scheduled clinical visits, 97.4% of planned plasma collections, 80.7% of stool samples, and all tumor biopsies. Wearable devices captured activity, heart rate, sleep, and blood oxygen saturation data during 95.0%, 84.2%, 90.6%, and 70.7% of total patient-days, respectively. Biospecimens met predefined quality control metrics across all molecular modalities. Engagement with mobile applications for pain and emotion reporting exceeded 80%.

**CONCLUSION:** The HDO study demonstrates the feasibility of comprehensive, longitudinal, multi-modal data collection in women with metastatic cancer. This protocol establishes an integrated framework for future analyses aimed at characterizing disease trajectories, defining molecular and physiological determinants of outcomes, and developing patient-specific computational models.

## Introduction

Precision oncology has transformed cancer care by matching treatments to tumor biomarkers, yet the predictive performance of genomics alone remains incomplete. Clinical benefit emerges from a complex interaction between tumor biology and the host’s physiological, behavioral, environmental, and treatment context. In metastatic disease, where trajectories are dynamic and treatment lines change over time, understanding longitudinal determinants of outcomes requires measurement frameworks that go beyond episodic clinic visits and single time point molecular profiling.

Multiple lines of evidence support the relevance of non tumor and non genomic dimensions for cancer outcomes, including—but not limited to—host inflammatory states, comorbidities and concomitant medications, lifestyle, and environmental exposures. At the same time, the widespread adoption of smartphones, wearable sensors, and digital PRO platforms makes it technically possible to capture continuous, low burden signals of physiology and behavior alongside scheduled clinical and molecular assessments. These developments enable a “high definition” approach to patient characterization, aligned with the broader paradigm of high definition medicine^2^, in which dense, longitudinal, multi modal data can complement standard oncology endpoints to better describe disease course and patient well being in real world care.

Large initiatives have demonstrated that deep phenotyping at scale is feasible in the general population, including resources such as UK Biobank^3^ and the All of Us Research Program^4^. In addition, precision health cohorts outside oncology have shown the scientific value of longitudinal multi omics and digital monitoring—e.g., integrative personal omics profiling (iPOP), wellness/behavioral coaching studies, “ageotype” longitudinal profiling, and deep phenotyping cohorts integrating clinical data with continuous monitoring and multi omics^5–9^. However, these frameworks are not designed around the specific operational constraints of advanced cancer patients oncology, where symptom burden, treatment toxicity, acute events, and time sensitive therapeutic decisions can challenge sustained adherence and biospecimen quality.

Within oncology, major efforts have achieved either depth in tumor molecular characterization or scale in clinicogenomic aggregation, but typically not integrated, prospective, synchronized capture of tumor/host multi omics, standardized longitudinal clinical phenotyping, and continuous digital monitoring. For example, TCGA established foundational multi omics tumor atlases largely from primary tumors; CPTAC expanded proteogenomic characterization; and AACR GENIE aggregates clinicogenomic data across institutions^10–13^. Longitudinal and metastatic programs such as TRACERx (lung cancer evolution)^14^, AURORA (metastatic breast cancer)^15^, and ITOMIC (intensive longitudinal multi site metastatic sampling)^16^ have advanced our understanding of tumor evolution and feasibility of repeated tissue profiling. Yet, to our knowledge, no multi center study in metastatic cancer has prospectively combined: (i) harmonized prospective clinical follow up with a dedicated electronic CRF, (ii) repeated blood and stool collection for multi omics across treatment, and (iii) continuous, remote, patient centric digital phenotyping (wearable + smartphone passive streams) plus scheduled PROs—at the intended depth and cadence—while explicitly treating feasibility and missingness as primary design constraints.

We launched the High Definition Oncology (HDO) observational study to address this gap. HDO profiles 300 women with treatment naïve advanced/metastatic breast, lung, or colorectal cancer across multiple centers in Spain (ClinicalTrials.gov: NCT06590506). The study integrates 11 longitudinal data modalities, spanning structured clinical phenotyping (EHR derived variables captured in a bespoke CRF), imaging, patient reported outcomes, continuous passive digital monitoring (wearable and smartphone), and multi omics (tumor whole exome sequencing; germline genotyping; longitudinal blood DNA methylation; gut metagenomics; plasma proteomics; plasma metabolomics; and stool metabolomics/exposome). By restricting enrollment to women, the study reduces sex related heterogeneity in physiology and behavior while enabling cancer specific and cross cancer analyses within common tumor types in females. The long term scientific objectives of HDO are to enable analyses that require synchronized longitudinal measurement across clinical, molecular and digital domains, including: (i) early prediction of functional deterioration and toxicity using combined physiological signals and PROs; (ii) linking longitudinal molecular dynamics (e.g., blood derived omics) to clinically meaningful events (progression, treatment changes); (iii) characterizing missingness mechanisms and their impact on inference in high burden oncology cohorts; and (iv) identifying longitudinal patient subtypes defined by trajectories rather than cross sectional states. These aims are explicitly downstream and hypothesis generating; they require mature follow up and larger sample sizes than an early accrual analysis.

A central uncertainty—and a critical barrier for the field—is whether a high burden, multi modal, longitudinal oncology study can be executed at scale without unacceptable loss of data completeness, biospecimen quality, or participant retention. This feasibility question is not trivial: adherence to ePRO and wearable monitoring varies widely across oncology studies, and high missingness can undermine both clinical interpretability and statistical validity. Therefore, HDO includes a pre specified internal pilot in the first 30 participants (10% of planned accrual), with formal progression criteria and a decision framework (Go / Go with amendments / No Go) as recommended for internal pilot designs^17^. Thresholds and monitoring procedures were pre specified in the study Statistical Analysis Plan and informed by published experience with electronic symptom monitoring and PRO implementation in oncology^18,19^ and by systematic reviews summarizing wearable devices adherence in cancer treatment trials^20^. For biospecimens, feasibility criteria incorporate quality and reporting expectations aligned with established biospecimen reporting and preanalytical coding frameworks and best practice guidance^21–23^.

## Results

### Study design

This is a prospective, observational study involving 9 Spanish hospitals (Institut Catala d’Oncologia (ICO)-Institut d’Investigació Biomèdica de Bellvitge; Hospital Virgen de la Macarena; University Hospital A Coruña (XXIAC-SERGAS); Hospital Son Espases; Hospital Universitario La Princesa; Hospital Universitario de Fuenlabrada; Hospital Universitario de Navarra; Hospital Clínico Universitario de Valencia; Hospital San Pedro Alcántara de Cáceres) and 3 research institutions (Centro Nacional de Investigaciones Oncologicas (CNIO); University Charles III of Madrid (UC3M) ; Polytechnical University of Madrid (UPM)) which aims to recruit a total of 300 female patients with advanced breast (BC; N=100), lung (LC; N=100) or colorectal (CRC; N=100) cancers. This target sample size was determined based on the need to ensure feasibility across multiple centers while allowing for statistically meaningful molecular and clinical subgroup analyses within each cancer type. Given the resource-intensive nature of the multi-omics profiling and longitudinal clinical monitoring, this cohort size represents a balance between comprehensive data depth and practical execution. The first patient was enrolled in February 2023 and the recruitment and monitoring will continue until the last patient’s demise. The study is coordinated by the Breast Cancer Clinical Research Unit (BCCRU) at CNIO, supported by two data analysis and artificial intelligence research groups at the UC3M and UPM and a Patient Advocacy Organization (CRIS Contra el Cancer).

### Recruitment and study adherence

Between February 16, 2023, and October 13, 2023, we recruited 30 patients. A 10% accrual rate was reached 239 days after the trial commenced. Initially, the trial began at a single hospital, with three additional study sites joining after three months, followed by two more at the four-month mark, another one after 5 months and the last one at the nine-month mark (October 2023). Nine months after the kick-off, a total of nine hospitals were participating. As of February 2026, the current accrual rate stands at 5.7 patients per month.

These first 30 patients were 13 breast, 13 lung and 4 colorectal metastatic cancer cases, with a mean age of 59.8 years (median 58.5 years, range 28-76 years). The mean recruitment pace, computed as the average number of new patients per month corrected by fraction of sites operative that month, was of 4.27 patients per month. At the 12-month mark (study report data cut-off date), these patients had been enrolled for a median of 217 days (range 29-365 days). One patient voluntarily withdrew consent (70 days) and 2 patients died after 29 and 181 days on trial, respectively (Figure 1).

**Figure 1.**
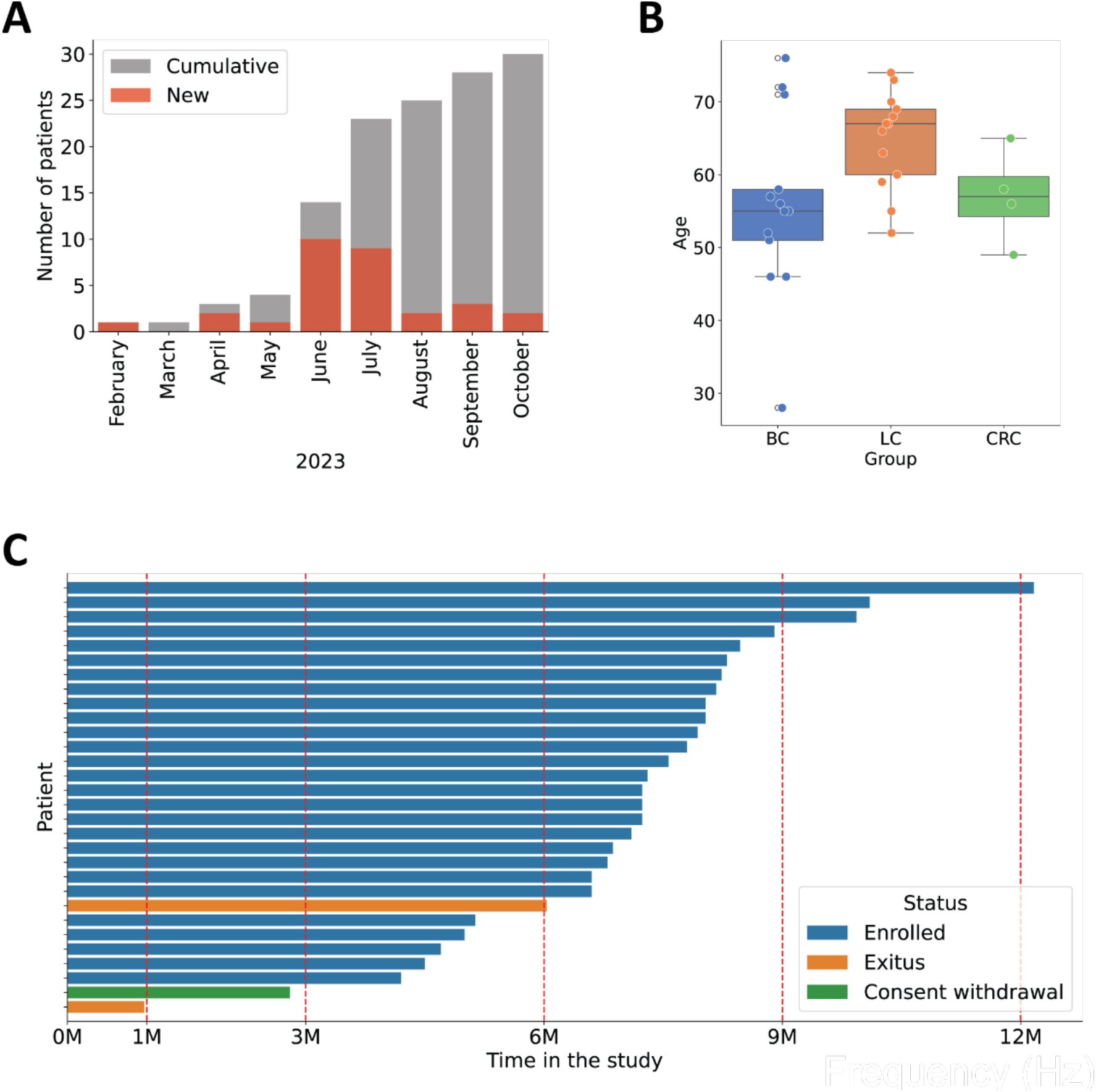
Overview of patient enrollment, demographics, and participation status in the study. (**A**) Monthly and cumulative patient enrollment over time from February to October 2023. Bars indicate the number of newly enrolled patients each month (red) and the cumulative total (gray). (**B**) Age distribution of participants by diagnostic group: breast cancer (BC), lung cancer (LC), and colorectal cancer (CRC). Boxplots show the median, interquartile range, and individual data points. (**C**) Patient retention and study participation timeline. Each horizontal bar represents an individual patient’s duration in the study. Bars are color-coded by status: actively enrolled (blue), deceased (exitus, orange), or withdrawn consent (green). Red dashed lines indicate key time points at first month and every 3-month intervals.

### Data and sampling completeness

All patients completed 100% of their due clinical visits. During these visits, 98.8% of the clinical laboratory tests, 100% of the medical history updates, 94.5% of social evaluation parameters and 94.05% of clinical evaluations (radiological evaluation, events -toxicty, treatment changes…-, ECOG) were completed. Out of all the planned samples, 97.37% of plasma, 80.7% of stool and 100% of the tumor samples were collected at the hospitals and shipped to the central laboratory (Figure 2A). Patients filled 81.14% of the planned questionnaires (Figure 2B); 28 patients reported pain values through the mobile app, with an average of 1.02 values per day (median 1, range 1-6). Similarly, out of the 30 patients, 28 imputed emotions through the eB2 application, with an average of 1.03 emotions per day (median 1, range 1-5) (Figure 2C).

**Figure 2.**
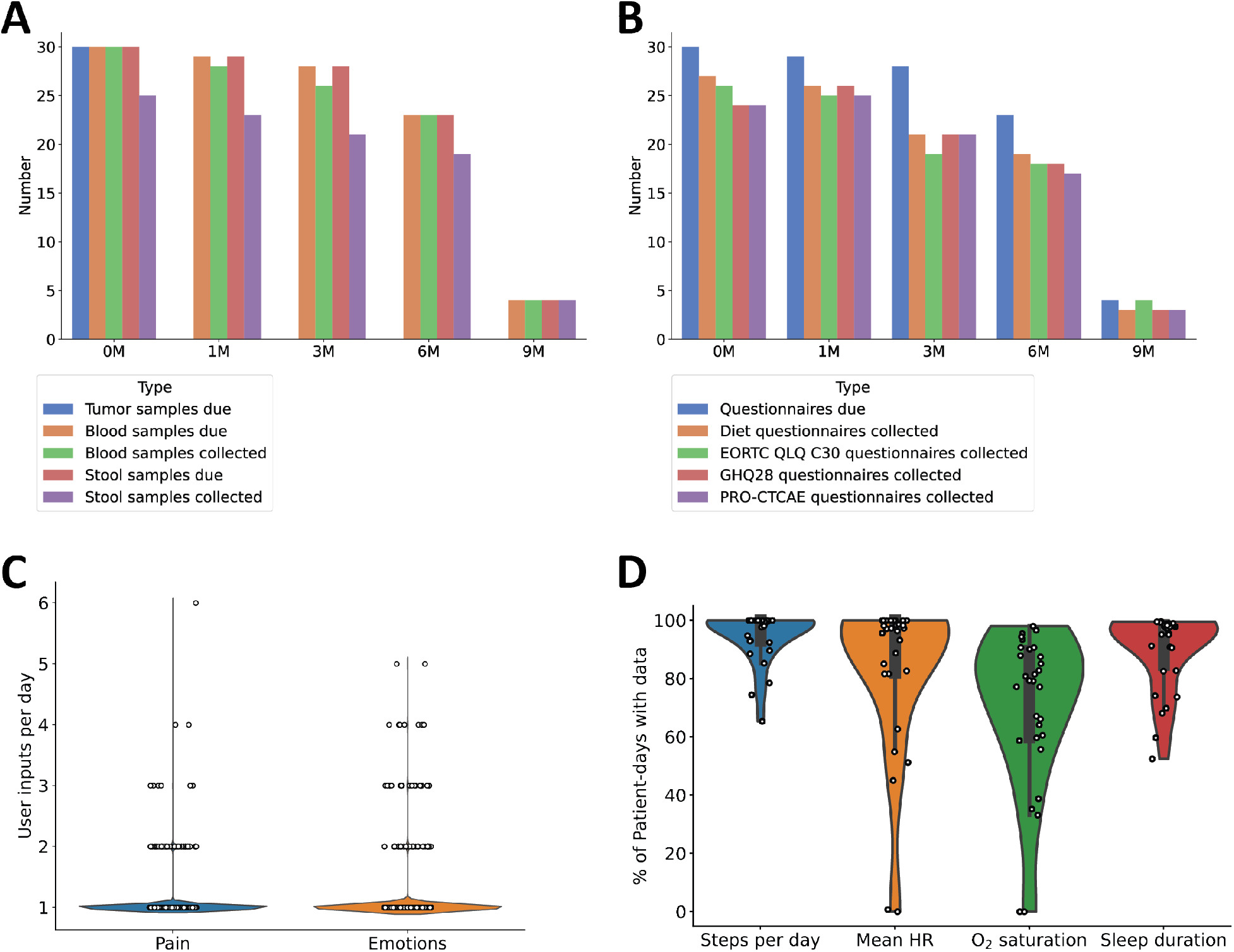
Data collection and digital health engagement. (**A**) Longitudinal sample collection overview showing the number of tumor, blood, and stool samples due and collected at baseline (0M) and at 1, 3, 6, and 9 months. (**B**) Questionnaires completed over time, displaying the number of questionnaires due and collected, including diet, EORTC QLQ-C30, GHQ28, and PRO-CTCAE questionnaires at the same intervals as in **A**. (**C**) Distribution of daily user inputs related to pain and emotions. Each dot represents a patient on a specific date. (**D**) Distribution of the percentage of patient-days with available data for physiological monitoring metrics: steps per day, mean heart rate (HR), oxygen (O_2_) saturation, and sleep duration. Each dot represents an individual patient.

Physiological monitoring captured activity (95.01%), heart rate (84.16%), sleep (90.64%), and blood oxygen saturation (70.73%) over total patient-days (Figure 2D). The observed variation in data capture rates reflects both technical and user-related factors. Activity data, based on accelerometry, demonstrated the highest capture rate due to its robustness and lower sensitivity to wear conditions. In contrast, heart rate and sleep data rely on continuous optical sensing, which may be affected by intermittent wear, improper device positioning, or motion artifacts. The notably lower capture rate for blood oxygen saturation may be attributed to its greater susceptibility to signal noise, the requirement for stable conditions (e.g., during sleep), and potential limitations in sensor performance. These discrepancies may also be influenced by device synchronization issues, user compliance, and periods of non-wear.

### Data Quality

All (100%) the biological samples collected at the central laboratory met the pre-established quality control criteria: a minimum concentration of 100 ng of DNA for tumor sample DNA WES, a minimum of 250 ng for genotyping and DNA methylation profiling, and a minimum of 100 ng for metagenomics sequencing. Tumor, blood and stool samples yielded 11.9 (0.29-70), 10.2 (0.4-59) and 9.0 (1.5-27) ug of DNA, respectively.

The total ion current, the median ion current and the coefficient of variation indicated that also all the plasma and stool samples were suitable for metabolomics profiling.

### Internal pilot feasibility outcomes and progression decision (Go/Amend/No-Go)

Feasibility was evaluated in the prespecified internal pilot (first 30 participants; 10% of planned accrual) by benchmarking observed performance against a priori progression criteria defined in the Statistical Analysis Plan (SAP; supplementary data). As recommended for internal pilot designs using progression criteria, feasibility endpoints were operationalized across five domains and classified using a traffic-light approach (RAG: Green/Amber/Red) to support a transparent scale-up decision: (i) recruitment and cohort representativeness; (ii) retention and longitudinal continuity; (iii) multimodal data and sampling completeness (scheduled core modalities and continuous digital streams assessed separately); (iv) biospecimen usability and quality control pass rates across molecular assays; and (v) data pipeline readiness, including acquisition-to-availability latency and critical error rates.

In brief, the internal pilot demonstrated: (1) high adherence to scheduled clinical follow-up and core assessments, (2) high completeness and QC pass rates for biospecimen collection and downstream molecular profiling, and (3) sustained capture of continuous digital streams with modality-specific completeness within prespecified expectations (see sections above). When assessed against the SAP progression criteria, 4 critical feasibility domains met Green thresholds and no critical domain met Red thresholds (Figure 3). Based on the prespecified decision rules, the internal pilot therefore resulted in an overall progression decision of [GO / GO-with-amendments] to proceed to full accrual. Amber domains, where present, triggered targeted corrective actions, including e.g., reinforced onboarding/troubleshooting SOPs; scheduling refinements; device-sync monitoring escalation, as documented in the SAP and protocol addendum (supplementary data).

**Figure 3.**
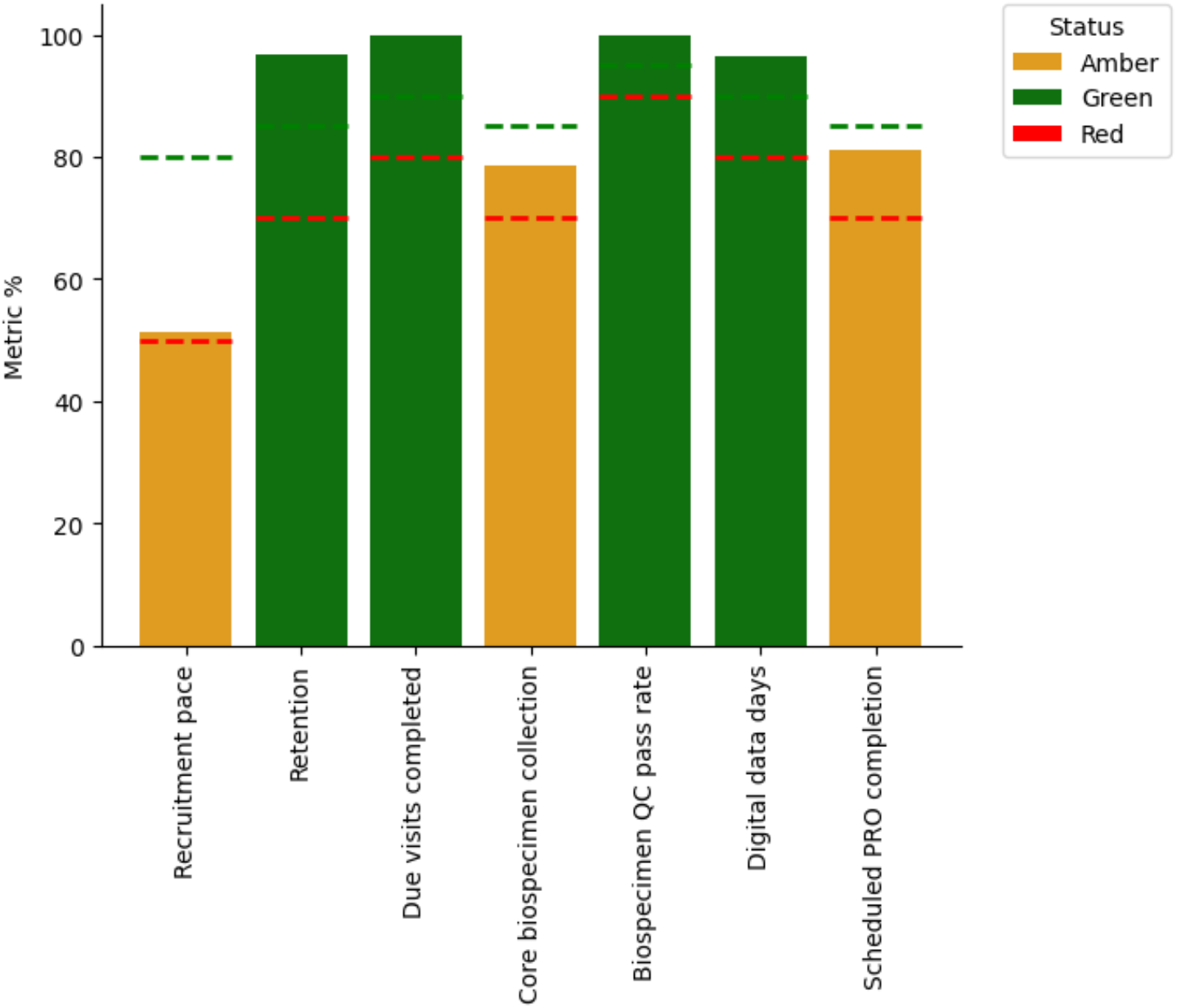
Evaluation of critical SAP progression criteria. Each bar indicates the percentage value of the corresponding critical SAP metric, with threshold values for Green and Red status indicated by dashed lines.

Following completion of the internal pilot and implementation of the above refinements, the study transitioned to full accrual. As of February 2026, (current submission data cut), a total of 205 participants have been enrolled across 9 sites, corresponding to a mean accrual rate of 5.7 participants per month. The extended recruitment curve illustrates scale-up after staged site activation and supports the feasibility of maintaining high-fidelity multimodal capture during continued enrollment (Supplementary figure **1**).

## Discussion

While genommic profiling has revolutionized targeted therapies, its predictive power remains incomplete, as highlighted by the substantial fraction of patients who fail to benefit from genomically matched treatments.^24–28^. Together with growing evidence that non-genetic factors (e.g., host physiology, microbiome, metabolic state, environmental exposures, lifestyle behaviors, and concomitant medications) contribute to cancer outcomes,^29–38^ these limitations motivate approaches that integrate tumor biology with longitudinal characterization of the host and its context.

The High-Definition Oncology (HDO) study was designed to address this implementation gap by prospectively synchronizing clinical phenotyping, repeated biospecimen collection for multiomics, and continuous patient-centric digital monitoring in women with metastatic breast, lung, or colorectal cancer. Although conceptually aligned with high-definition medicine^2^, the primary contribution of this manuscript is operational and methodological: determining whether a high-burden, multi-modal, longitudinal profiling framework can be executed at scale in routine metastatic oncology care with acceptable adherence, completeness, and biospecimen usability.

A key advance in the revised design is that feasibility was prespecified and evaluated as an internal pilot embedded within the study. Following recommendations for progression criteria in internal pilots^17^, feasibility was operationalized across critical domains (recruitment/representativeness, retention/longitudinal continuity, multimodal completeness, biospecimen QC pass rates, and data pipeline readiness) and assessed using a traffic-light (Red/Amber/Green) framework defined a priori in the Statistical Analysis Plan. In the first 30 consecutively enrolled participants (10% of planned accrual), the study met prespecified critical progression criteria, resulting in an overall Go decision to proceed to full accrual. This decision was supported by complete attendance of scheduled clinical visits, high completion of core clinical phenotyping and good completion of scheduled questionnaires, high overall completeness across biospecimens (with lower completeness for stool than plasma), sustained capture of wearable-derived streams across total patient-days (steps, heart rate, sleep, and nocturnal oxygen saturation) with expected stream-level variability, and 100% QC pass rates for collected biospecimens across molecular modalities. Together, these findings show that dense, synchronized capture across heterogeneous data streams is achievable in a clinically unstable metastatic population—an uncertainty that often limits adoption of deeply phenotyped oncology cohorts. Following the Go decision, accrual proceeded to full enrollment under the same standardized framework.

Beyond feasibility per se, the internal pilot provides quantitative benchmarks that can inform the design of future high-burden cohorts and trials. When missingness is substantial, interpretability and statistical validity can be compromised; early evidence on achievable completion rates and biospecimen usability helps teams calibrate realistic study intensity and staffing needs. Importantly, this report is not intended to draw clinical conclusions or validate predictive models. Rather, it establishes the upstream measurement fidelity required for downstream analyses. As the cohort matures, HDO is designed to enable hypothesis-generating analyses that require synchronized longitudinal measurement, including: (i) early prediction of functional deterioration and treatment-related toxicity using integrated physiological signals and PROs; (ii) linking longitudinal molecular dynamics (e.g., blood-derived omics) to clinically meaningful events such as progression and treatment modifications; (iii) characterizing missingness mechanisms and their impact on inference in high-burden oncology cohorts; and (iv) identifying longitudinal patient subtypes defined by trajectories rather than cross-sectional states.

The operational success observed in the internal pilot likely reflects both participant engagement and the prospective implementation framework deployed from study launch. Because these procedures were implemented throughout enrollment, their individual causal contribution cannot be isolated; nevertheless, they constitute a practical blueprint that other teams can replicate. Key elements (Box 1) include standardized pretreatment onboarding and device/app training, a single-vendor wearable strategy to reduce heterogeneity, centralized dashboards with proactive troubleshooting triggers (e.g., response to ≥48-hour data gaps), alignment of PRO administration with clinic workflows, harmonized biospecimen SOPs with centralized QC gates, staged multicenter activation with continuous feedback, and privacy-by-design governance. Collectively, these measures address common failure modes in digital and biospecimen-intensive studies— device non-wear and syncing issues, inconsistent preanalytical handling, heterogeneous capture definitions across sites, and delayed data availability.

Positioning HDO within the broader landscape clarifies why this feasibility evidence matters. Large population initiatives such as UK Biobank and the All of Us Research Program demonstrate the feasibility and value of deep phenotyping at scale^3,4^, and precision-health cohorts outside oncology have shown the potential of combining longitudinal multi-omics with dense phenotyping ^5–9^. Within oncology, major resources have achieved either tumor molecular depth or clinicogenomic scale (e.g., TCGA^39^, CPTAC^12^) or clinicogenomic scale (GENIE^13^) and longitudinal programs such as TRACERx, AURORA and ITOMIC have advanced repeated sampling and evolutionary analyses in selected contexts^141516^. However, metastatic oncology remains under-served by cohorts that prospectively synchronize harmonized clinical phenotyping (including a bespoke CRF), repeated host/tumor multi-omics, continuous digital phenotyping, and scheduled PROs at high cadence across multiple centers, while explicitly treating feasibility and operational scalability as primary design constraints. To avoid misleading cross-study quantitative comparisons given heterogeneity in denominators and capture definitions, we provide a modality mapping of representative initiatives in Table 1

**Table 1.** Summary of representative studies in precision oncology. *Via TCIA (WSI, and Radiology),**Not performed systematically for all subjects, ***Clinical follow up and outcomes are recorded, but no longitudinal sampling.

| Study | Focus | N | Cancer types | Longitudinal design | Prospective harmonized clinical CRF | Imaging | PROs | Digital phenotyping phone | Digital phenotyping watch | Tumor genomics | Tumor transcriptomics | Tumor proteomics | Germline genomics | Germline epigenomics | Blood transcriptomics | Plasma metabolome | Plasma proteome | Stool metagenomics | Exposome characterization |
| --- | --- | --- | --- | --- | --- | --- | --- | --- | --- | --- | --- | --- | --- | --- | --- | --- | --- | --- | --- |
| TCGA | Molecular characterization of primary tumors | 11160 | Pan-cancer (33 types) | NO | NO | YES* | NO | NO | NO | YES | YES | YES | YES | YES** | YES** | NO | NO | NO | NO |
| CPTAC | Tumoral proteo-genomics | >1500 | Pan-cancer (10 types) | NO | NO | YES* | NO | NO | NO | YES | YES | YES | YES | YES** | YES** | NO | NO | NO | NO |
| GENIE | Clinical and genomic register of clinical practice | 227696 (GENIE 19.0, January 2026) | Pan-cancer (80+ types) | NO*** | NO | NO | NO | NO | NO | YES | NO | NO | NO | NO | NO | NO | NO | NO | NO |
| TRACERx | Clonal evolution and longitudinal intratumoral heterogeneity | 428 | Early stage NSCLC | YES | YES | YES | NO | NO | NO | YES | YES | NO | YES | NO | NO | NO | NO | NO | NO |
| AURORA | Biology and evolution of metastatic breast cancer | 1300, extended with 252 additional patients | Metastatic breast cancer | YES | YES | YES | YES | NO | NO | YES | YES | NO | YES | NO | YES | NO | NO | NO | NO |
| ITOMIC | Multi-omic profiling and digital phenotyping | 31 | Advanced solid tumors (pan-cancer) | YES | YES | YES | YES | YES | YES | YES | YES | YES | YES | YES | YES | YES | YES | YES | YES |
| HDO | Multi-modal longitudinal profiling of advanced cancers | 300 | Metastatic breast, lung and colorectal cancers | YES | YES | YES | YES | YES | YES | YES | YES | YES | YES | YES | YES | YES | YES | YES | YES |

### Box 1.

**HDO Operational Framework**

1. **Standardized pretreatment onboarding:** dedicated training visit (Day −14 to 0) to install the study app, provision the wearable, verify permissions/sync, and rehearse patient tasks.
2. **Single-vendor wearable across sites:** one device model to reduce heterogeneity in sensing, patient instructions, and troubleshooting.
3. **Centralized monitoring dashboard with proactive triggers:** coordinators monitor stream continuity and contact participants when ≥48 hours without incoming data are detected to resolve charging/sync/permission issues.
4. **Burden-aware charging instructions:** participants are instructed to charge the wearable during daytime to preserve nocturnal sensing (critical for sleep and SpO_2_ capture).
5. **Redundant PRO capture aligned with visits:** questionnaires are available in-app and scheduled to coincide with hospital visits whenever possible to minimize missed assessments.
6. **Harmonized biospecimen SOPs and centralized QC gates:** standardized collection/processing/shipping procedures across sites with predefined QC acceptance criteria at the central laboratory to avoid unusable -omics datasets.
7. **Staged multicenter ramp-up with continuous feedback:** site activation in waves plus continuous feedback loops to reduce inter-site variability over time.
8. **Governance and privacy-by-design:** pseudonymization, role-based access, and GDPR-aligned governance enabling cross-center operations and secure data flows.

Several limitations should be considered. First, internal pilot findings derive from a limited early-accrual cohort and follow-up window and may not capture late-emerging adherence challenges. Second, participation requires smartphone access and sufficient digital literacy, and the cohort is restricted to women; these constraints may limit generalizability to other populations and settings. Third, feasibility performance depends on the specific operational infrastructure and vendor ecosystem used here, and results may differ with alternative platforms or in health systems with different staffing resources. Finally, as an observational cohort, HDO is not designed to establish causal effects; downstream analyses will require careful handling of confounding, informative censoring, and missingness.

In summary, HDO demonstrates—using prespecified progression criteria and a Go decision framework—that comprehensive, longitudinal, multimodal profiling is operationally feasible in women with metastatic cancer across multiple centers, with high adherence, biospecimen usability, and sustained digital capture. By providing quantitative feasibility benchmarks and a replicable implementation blueprint, this protocol and internal pilot report lays the foundation for future high-definition oncology cohorts and for integrative analyses linking molecular, physiological, and behavioral dynamics to clinically meaningful outcomes.

## Supporting information

Statistical Analysis Plan

## Data Availability

All data produced in the present study are available upon reasonable request to the authors

