## Supplementary material for "Longitudinal clinical, physiological and molecular profiling of female metastatic cancer patients: protocol and feasibility of a multicenter high-definition oncology study": Statistical Analysis Plan

**High-Definition Oncology (HDO)
Statistical Analysis Plan (SAP)**

Protocol Attachment – Internal Pilot Feasibility Progression Criteria

Version: v1.0
Date: 2023-04-05
ClinicalTrials.gov: NCT06590506

### Table of Contents

1. Purpose and scope
2. Study overview
3. Objectives and endpoints
4. Analysis populations and data cut
5. General statistical principles
6. Descriptive analyses for the mature cohort
7. Internal pilot feasibility evaluation (Go/Amend/No-Go)
8. Missing data and data quality
9. Reporting conventions
10. Deviations and amendments
Appendices: Schedule of assessments; Variable dictionary; Feasibility criteria table; Abbreviations; References

### 1. Purpose and scope

This Statistical Analysis Plan (SAP) specifies the planned analyses for the High-Definition Oncology (HDO) observational cohort study. The SAP is intended as a protocol attachment to (i) standardize routine descriptive analyses across modalities, (ii) prespecify an internal pilot feasibility assessment with progression criteria to support a Go/Amend/No-Go decision for scale-up to the full intended sample size, and (iii) promote transparency and reproducibility across multiple downstream manuscripts.

### 2. Study overview

HDO is a prospective, multicenter observational cohort conducted in Spain. Eligible participants are adult women with newly diagnosed, non-curable locally advanced or metastatic breast, lung, or colorectal cancer who are treatment-naïve for the advanced/metastatic setting. Participants undergo longitudinal multimodal profiling including: (a) clinical and treatment data captured in a dedicated eCRF/EHR abstraction; (b) scheduled biospecimen collection (blood and stool) and archival tumor tissue retrieval for multi-omic analyses; (c) imaging (DICOM archiving for future radiomics); and (d) continuous/passive physiologic and behavioral monitoring using a smartphone application and a standardized wearable device.

The date of first systemic treatment dose is considered Day 1. Core follow-up procedures and sample collection are repeated at Month 1, Month 3, and then every 3 months until disease progression, death, or voluntary withdrawal. Digital monitoring is continuous.

### 3. Objectives and endpoints

#### 3.1 Primary scientific objectives (mature cohort)

The primary scientific objective of HDO is to build longitudinal, multimodal patient profiles that capture host state and disease trajectory, and to evaluate associations between profile dynamics and clinically relevant outcomes. Because HDO is observational and hypothesis-generating, downstream manuscripts may specify additional endpoints and hypotheses; however, core outcome definitions and baseline descriptive analyses are prespecified herein.

#### 3.2 Core clinical endpoints (definitions)

Primary time-to-event endpoint: Progression-free survival (PFS), defined as time from Day 1 to first documented disease progression (radiographic or clinical, per treating physician assessment and standard practice) or death from any cause, whichever occurs first. Patients without an event will be censored at last adequate disease assessment date.

Secondary clinical endpoints may include overall survival (OS), time to treatment discontinuation, objective response (when measurable disease is present), and patient-reported outcome (PRO) trajectories, as defined in protocol and the data dictionary.

#### 3.3 Internal pilot feasibility objective

A prespecified internal pilot will evaluate feasibility of recruitment and high-fidelity, longitudinal multimodal data capture. Feasibility will be assessed in the first 30 consecutively enrolled eligible participants (the 'Pilot Set'), using prespecified progression criteria (Green/Amber/Red). The feasibility evaluation supports a Go/Amend/No-Go decision for scaling to the full target sample size.

#### 3.4 Feasibility endpoints (Pilot Set)

Feasibility endpoints are grouped into: (i) recruitment and retention; (ii) completion of scheduled clinical visits and eCRF/EHR variables; (iii) biospecimen collection completeness and laboratory QC pass rates; (iv) digital monitoring completeness (valid days) across core streams; (v) PRO completion rates; and (vi) data pipeline readiness (time-to-ingestion, error rates). Operational definitions are provided in Section 7 and Appendix A.

### 4. Analysis populations and data cut

#### 4.1 Analysis sets

Enrolled Set: all consented participants registered in the study database.
Onboarded Set: enrolled participants who complete digital onboarding (app installed and wearable linked) and initiate monitoring.
Pilot Set: the first 30 consecutively enrolled eligible participants with monitoring initiated (subset of the Onboarded Set).
Mature Cohort Set: all participants accrued to the full target sample size, analyzed according to the data cut specified per manuscript.

#### 4.2 Data cut and database lock

For the Pilot Set feasibility evaluation, a fixed data cut will be used: each pilot participant will be followed from Day −14 to Day +90 (or until earlier progression, death, or voluntary withdrawal). Pilot feasibility analyses will be performed after all 30 participants reach the Day +90 window or an earlier censoring event, and after completion of data quality checks and database lock.

### 5. General statistical principles

This SAP emphasizes estimation over hypothesis testing. Unless explicitly stated, analyses will be descriptive with appropriate measures of uncertainty (95% confidence intervals [CI]). Two-sided tests with α=0.05 may be used for prespecified comparisons in downstream manuscripts; multiplicity control will be described per manuscript depending on the number of endpoints and models.

Continuous variables will be summarized as mean (SD) or median (IQR) depending on distribution. Categorical variables will be summarized as counts (%). Time-to-event endpoints will be summarized by Kaplan–Meier curves with median estimates and 95% CI.

#### 5.1 Software and reproducibility

Analyses will be performed in Python and/or R. Scripts will be version-controlled and archived with the data cut used for each analysis. Random operations (e.g., matching) will use fixed seeds.

### 6. Descriptive analyses for the mature cohort

The following descriptive analyses will be produced routinely for each data cut and may be reused across manuscripts:

• Baseline cohort description: demographics, tumor type, stage, key clinical variables, comorbidities, ECOG, and baseline labs.
• Follow-up and attrition: follow-up time distribution; reasons for withdrawal; censoring patterns.
• Clinical outcomes: Kaplan–Meier for PFS and OS overall and by tumor type.
• Data completeness dashboard: per modality and per center (counts due vs collected; % complete; QC pass rates).
• Digital adherence: distributions of valid-day percentages per stream and over time; time-to-first gap analyses.
• PRO completion and trajectories: completion rates; longitudinal mixed models for PROs if used in manuscripts.

### 7. Internal pilot feasibility evaluation (Go/Amend/No-Go)

#### 7.1 Rationale and principles

The internal pilot is designed to determine whether the intended intensity of multimodal data capture is operationally scalable in women with metastatic cancer. Progression criteria are prespecified using a traffic-light approach: Green (Go), Amber (Go with amendments), Red (No-Go or major redesign). Criteria are applied to key feasibility domains and interpreted jointly to support a decision.

#### 7.2 Operational definitions

Scheduled clinical visits and biospecimen collections are considered 'due' according to the protocol schedule (baseline, Month 1, Month 3 within the pilot window). A visit or biospecimen is 'complete' if collected within the allowed window and meets minimum processing requirements.
Digital monitoring 'valid day' definitions for core streams:
• Steps: valid if steps are registered in ≥75% of non-sleep hourly intervals.
• Heart rate: valid if ≥50% of the day is covered by determinations.
• Sleep: valid if ≥3 hours of sleep is recorded.
• SpO2: valid if ≥1 nocturnal determination is recorded.
• GPS/location: valid if ≥50% day coverage of GPS signal.
These definitions may be refined in the data dictionary but will remain fixed within a given data cut.

#### 7.3 Feasibility metrics

Metrics will be computed both overall and stratified by center and tumor type where numbers allow.

Recruitment:
• Accrual rate per center (patients/month) and overall.
• Time to enroll the first 30 participants.

Retention:
• Proportion of Pilot Set participants remaining on study and providing any data through Day +90 (or to earlier event).
• Reasons for withdrawal.

Clinical visit and eCRF completeness:
• Proportion of due visits completed within window.
• Proportion of core eCRF variables complete at each visit.

Biospecimens:
• Proportion of due blood and stool collections successfully obtained.
• QC pass rate for each biospecimen type/assay (e.g., minimum yield, integrity metrics).

Digital monitoring:
• Any-data days: proportion of observed patient-days with ≥1 digital parameter received.
• Stream-specific valid-day percentages per participant for core streams.
• Time to first ≥48-hour gap per stream.

PROs:
• Completion rate of scheduled questionnaires (e.g., EORTC QLQ-C30, GHQ-28).
• Completion rate of daily VAS pain entries (if required) and optional entries (emotions) reported separately.

Data pipeline readiness:
• Median time from acquisition to ingestion into the analytic warehouse.
• Proportion of records with critical errors requiring manual resolution.

#### 7.4 Prespecified progression criteria (traffic light)

Table 1 (Appendix B) defines prespecified thresholds for each metric. Criteria are categorized as Critical (must not be Red) or Supportive (inform amendments). Thresholds are informed by prior feasibility/internal pilot methodology and by published adherence ranges for ePRO and wearable monitoring and biospecimen QA standards.

#### Table 1. Internal pilot progression criteria (summary)

| Domain / Metric | Denominator | Green (Go) | Amber (Amend) | Red (No-Go) | Critical? |
| --- | --- | --- | --- | --- | --- |
| Recruitment pace (overall) | Planned accrual in pilot period | ≥80% of planned | 50–79% | <50% | Yes |
| Retention to Day +90 | Pilot Set participants | ≥85% | 70–84% | <70% | Yes |
| Due visits completed | All due visits in pilot window | ≥90% | 80–89% | <80% | Yes |
| Core biospecimen collection | All due tumor/blood/stool collections | ≥85% | 70–84% | <70% | Yes |
| Biospecimen QC pass rate | Collected biospecimens | ≥95% | 90–94% | <90% | Yes |
| Any digital data days | Observed patient-days | ≥90% | 80–89% | <80% | Yes |
| Valid days: HR/steps/sleep (median) | Per-participant % valid days | Median ≥80% | Median 65–79% | Median <65% | Supportive |
| Valid days: GPS/app-use (median) | Per-participant % valid days | Median ≥70% | Median 50–69% | Median <50% | Supportive |
| Scheduled PRO completion | All scheduled PRO prompts | ≥85% | 70–84% | <70% | Yes |
| Data ingestion latency | Core streams/records | ≥90% within ≤14 days | 70–89% within ≤14 days | <70% within ≤14 days | Supportive |

Notes: Thresholds may be refined during protocol amendment prior to pilot analysis, but must be finalized and version-controlled before database lock. For supportive metrics, Red triggers mandatory amendments but does not automatically imply No-Go if all critical criteria are Green/Amber.

#### 7.5 Decision algorithm

The overall decision will be based on the joint pattern of criteria:
• Go: all Critical criteria are Green, or at most one Critical criterion is Amber with a prespecified corrective action plan.
• Go with amendments: no Critical criterion is Red, and (i) ≥2 Critical criteria are Amber and/or ≥2 Supportive criteria are Red/Amber, requiring operational improvements before scaling.
• No-Go: any Critical criterion is Red, or a pattern of failure suggesting unresolvable burden (e.g., persistent low retention coupled with poor PRO and digital completeness).
The steering committee will document the decision and the rationale in a dated memorandum linked to the SAP version.

#### 7.6 Statistical methods for feasibility endpoints

For each feasibility proportion (e.g., retention, completion), the point estimate and 95% CI will be reported. Wilson score intervals will be used for binomial proportions. For accrual rates (counts per unit time), Poisson rate estimates with 95% CI will be reported.
Stream-level valid-day percentages will be summarized across participants using median (IQR) and visualized with boxplots/violin plots. Time-to-first ≥48-hour gap will be summarized using Kaplan–Meier methods.

Sensitivity analyses will include: (i) excluding participants who progressed or died within 30 days (to separate disease-related attrition); (ii) stratification by center and tumor type; (iii) repeating metrics using alternative but prespecified valid-day definitions if applicable.

### 8. Missing data and data quality

Missingness will be quantified and categorized by modality and reason (technical failure, participant choice, clinical event, logistical issues). For feasibility endpoints, missing data will generally be treated as non-completion unless a modality is not due (e.g., participant progresses before Month 3 visit). For scientific analyses, handling of missing data (e.g., mixed models, multiple imputation, model-based approaches) will be prespecified in downstream manuscript analysis plans.

### 9. Reporting conventions

Pilot feasibility results will be reported in a dedicated section including: (i) baseline characteristics of the Pilot Set; (ii) feasibility outcomes versus progression criteria (table); (iii) center-level dashboards; and (iv) decision statement (Go/Amend/No-Go). All figures and tables will specify denominators and observation windows.

### 10. Deviations and amendments

Any changes to the SAP after initiation of the pilot will be documented with version number, date, and justification. Changes affecting feasibility thresholds must be finalized prior to pilot database lock.

### Appendix A. Schedule of assessments (pilot window summary)

Day −14 to Day 0: onboarding, baseline clinical assessments, baseline biospecimens.
Day 1: treatment start.
Month 1 (±window): clinical assessment, labs, blood/stool.
Month 3 (±window): clinical assessment, labs, blood/stool (if within pilot window).
Month 6 (±window): clinical assessment, labs, blood/stool (if within pilot window).
Month 9 (±window): clinical assessment, labs, blood/stool (if within pilot window).
Month 12 (±window): clinical assessment, labs, blood/stool (if within pilot window).
Continuous: wearable and smartphone passive monitoring; daily VAS; scheduled PRO questionnaires.

### Appendix B. Detailed progression criteria table (editable template)

This appendix may be expanded to include per-modality criteria (e.g., per biospecimen type, per PRO instrument), center-specific thresholds, and prespecified corrective actions.

### Appendix C. Abbreviations

AUC: Area under the curve; CI: Confidence interval; eCRF: Electronic case report form; EHR: Electronic health record; HDO: High-Definition Oncology; HR: Heart rate; OS: Overall survival; PFS: Progression-free survival; PRO: Patient-reported outcome; QC: Quality control; SAP: Statistical analysis plan.

### Appendix D. References (key methodology)

1) Eldridge SM et al. CONSORT 2010 statement: extension to randomised pilot and feasibility trials. BMJ. 2016.
2) Avery KNL et al. Informing efficient randomised controlled trials: challenges in developing progression criteria for internal pilot studies. BMJ Open. 2017.
3) Mellor K et al. Recommendations for progression criteria in pilot trials. (as applicable).
4) Moore HM et al. Biospecimen Reporting for Improved Study Quality (BRISQ). Cancer Cytopathology. 2011.
5) ISBER. Standard PREanalytical Code (SPREC). Versions as applicable.
6) NCI Best Practices for Biospecimen Resources. 2016.
7) Basch E et al. PRO-TECT trial (ePRO adherence). 2025.
