## Supplementary Figures for "Longitudinal clinical, physiological and molecular profiling of female metastatic cancer patients: protocol and feasibility of a multicenter high-definition oncology study"

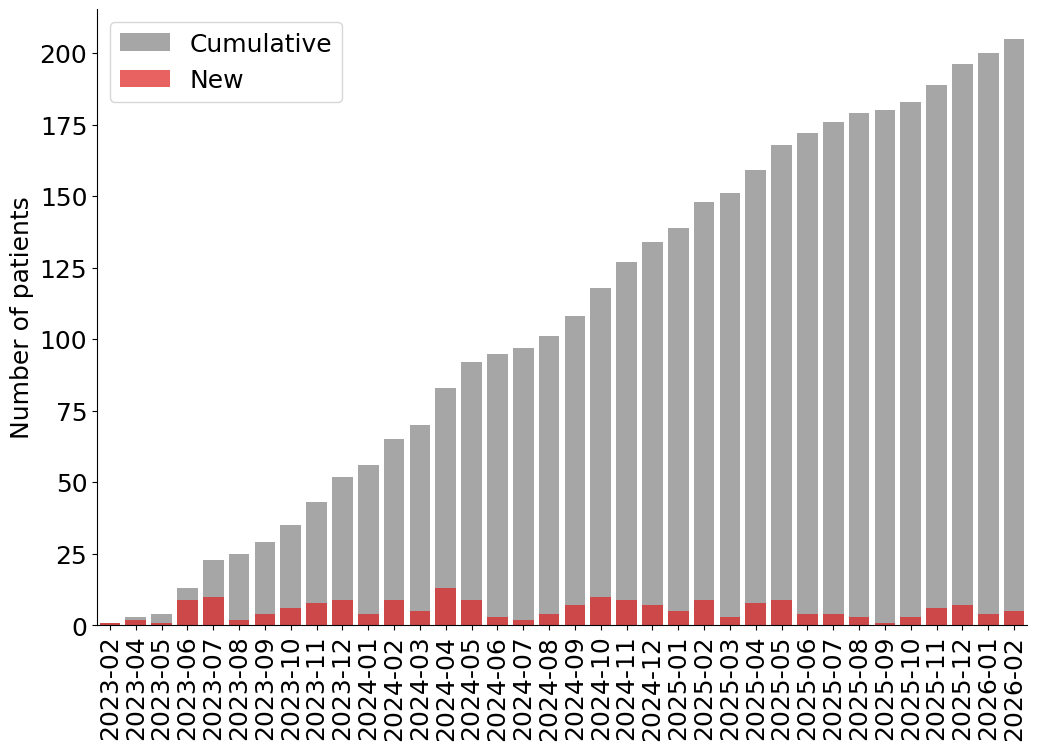

**Supplementary Figure 1. Evolution of patient enrollment.**

Monthly and cumulative patient enrollment over time from February 2023 to Februrary 2026. Bars indicate the number of newly enrolled patients each month (red) and the cumulative total (gray).
