## Supplementary Methods for "Longitudinal clinical, physiological and molecular profiling of female metastatic cancer patients: protocol and feasibility of a multicenter high-definition oncology study"

##### Accrual

The inclusion criteria for the study require participants to be female with a histologically confirmed diagnosis of oncological disease of solid origin (breast cancer (BC), lung cancer (LC), or colorectal cancer (CRC)) at stage III or IV, not amenable to curative treatment. Patients must be treatment-naïve for metastatic disease and candidates for first-line systemic therapy. Previous treatments for the metastatic setting such as palliative analgesic radiotherapy for bone metastases or cranial radiotherapy—whether for symptomatic brain metastases or at the investigator’s discretion—are permitted, as long as inclusion occurs after such symptoms are controlled.

For the BC cohort, patients must have hormone receptor-positive (HR+), HER2-negative tumors and be planned for treatment with a CDK4/6 inhibitor in combination with endocrine therapy. Patients in the LC and CRC cohorts may be enrolled regardless of tumor subtype, provided they are eligible for standard first-line treatment regimens. Inclusion is also permitted for patients participating in clinical trials, as long as the treatment is known and not part of a double-blind protocol. Acceptable examples include—but are not limited to—patients receiving investigational SERDs or CDK4/6 inhibitors in open-label studies, or those initiating triplet therapies that include chemotherapy and immunotherapy components.

All participants must have an ECOG performance status of less than 2 (i.e., 0 or 1), be capable of wearing a smartwatch, and possess sufficient digital literacy to operate a mobile phone, access email, and interact with the study’s designated applications. Patients must also be willing and able to complete electronic questionnaires covering quality of life, nutrition, and mental health domains. Informed consent must be obtained in accordance with ICH E6R2 guidelines prior to the initiation of any study-specific procedures.

To ensure real-world applicability, no restrictions are placed on comorbidities, co-medications, or life expectancy. Pregnant women are not excluded. However, patients are not eligible if they have previously received systemic therapy for metastatic disease, are enrolled in double-blind treatment trials, or have a current or past malignancy that could interfere with protocol compliance or data interpretation (excluding basal cell carcinoma of the skin or cervical carcinoma in situ, if treated curatively). Additionally, individuals with therapeutic electronic implants such as pacemakers, defibrillators, or cardiac resynchronizers are excluded due to potential interference with wearable devices. The inclusion and exclusion criteria are also detailed in the study's registration at ClinicalTrials.gov (Identifier: NCT06590506)^1^.

##### Patient training visit

Besides the engagement with serial tumor, blood and fecal sampling, a key aspect for study success is the extent to which the patients are familiar with the wearable and tracking devices required for this trial. Upon enrollment, each patient is assigned a unique numerical identifier and registered in a monitoring platform called eB2 MindCare^2^.

eB2 MindCare includes a patient-facing mobile application and a research dashboard for study coordinators and researchers. The application automatically collects a variety of data, including patients' emotions (inputted at will) and daily pain levels (prompted daily), and administers periodic health questionnaires (EORTC QLQ-30, GHQ28, PRO-CTCAE at baseline, month 1, and every 3 months thereafter, typically coinciding with hospital visits). Additionally, dietary data related to adherence to the Mediterranean diet is also collected periodically via the app. The MindCare application further gathers passive data such as phone usage patterns and activity derived from the smartphone's sensors. The necessary permissions are granted upon installation, allowing seamless and secure data integration directly with eB2 databases.

The monitoring via the eB2 MindCare app is complemented by physiological tracking using a wearable device (Vivosmart 4 activity tracker, Garmin, USA). Patients receive detailed instructions and documentation for operating the Garmin wearable and installing its companion app, Garmin Connect, on their smartphones. The Garmin wearable monitors physiological parameters continuously, including blood oxygen saturation during sleep periods. To minimize data gaps, patients are specifically instructed to recharge the device during daytime hours, ensuring continuous data collection overnight. Typically, the rechargeable battery lasts up to 5 days and requires less than 30 minutes for a full charge.

The Garmin Connect app transfers the wearable's physiological data to the eB2 MindCare platform, facilitating comprehensive physiological and behavioral monitoring. Study coordinators are instructed to promptly notify eB2 data managers if a patient experiences technical difficulties, changes or loses their phone, or uninstalls the app. This ensures quick resolution, preserving data integrity and study continuity.

Patients are informed of the study’s tentative schedule and their upcoming scheduled clinical visits.

##### Samples and data collection

The study aims to obtain clinical and molecular profiles from the patients at regular intervals from the diagnosis until disease progression or complete recovery. Patients are scheduled for clinical consultation and baseline sample and collection visit before the first planned treatment dose of their systemic treatment. Subsequent timepoints are 1 month after the first treatment dose, and every three months thereafter.

At the baseline visit, a tumor biopsy, blood sample, and stool sample are collected from the patient. Clinical data are then captured using a customized electronic case report form (CRF) designed specifically for the study, which includes 24 clinical fields, 18 demographic fields, and 96 bloodwork fields. These fields are populated by dedicated data entry staff, who gather information from the electronic health records (EHR), clinician input, and lab results from bloodwork and biochemistry. All CRF fields are completed during the baseline visit, while subsequent visits only involve updates to the clinical and bloodwork data.

Genomic profiles of the patients and their tumors, blood DNA methylation, gut microbiome, blood plasma proteome, blood plasma metabolome, and fecal metabolome are derived from the baseline samples. On subsequent visits, additional blood and stool samples are collected to generate further molecular data —blood DNA methylation, gut microbiome, blood plasma proteome, blood plasma metabolome, and fecal metabolome— (Figure 1).

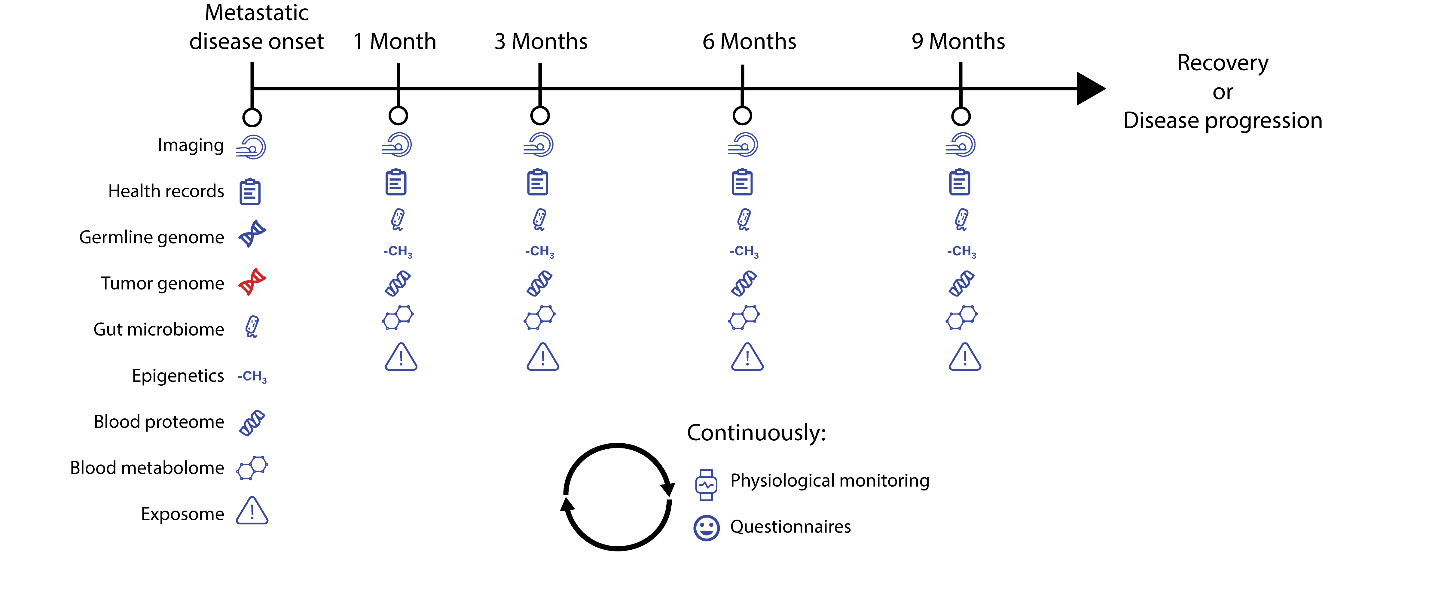

**Figure 1.** **Schematic overview of the longitudinal data collection strategy in a clinical trial following metastatic disease onset.**

Data types are collected at metastatic disease onset and at subsequent intervals of 1, 3, and every 3 months thereafter until recovery or disease progression. Data sources include imaging, health records, germline genome, tumor genome, gut microbiome, epigenetics, blood proteome, blood metabolome, and exposome. Germline and tumor genome data are collected only at baseline (metastatic disease onset), while other data types are collected repeatedly at designated time points. In parallel, physiological monitoring data and patient-reported outcomes are collected continuously. This multi-modal and longitudinal approach enables comprehensive characterization of disease trajectory and therapeutic outcomes.

##### Sample processing

Routine blood parameters are determined at the study site laboratories. A total of 4 4-ml EDTA tubes are collected for hematology analysis, while 1 tube of 3 ml venous blood is obtained for gasometry. A 4 ml coagulation-citrate tube is used for coagulation testing, and several 5 ml biochemistry tubes are collected for biochemical assays, including markers of renal, hepatic, and cardiovascular function. Further samples are collected at the study sites and sent to the central laboratory for additional analysis: two 10-ml BD Vacutainer CPT tubes are collected for peripheral blood mononuclear cell (PBMCs) extraction, and two 10-ml BD Vacutainer EDTA tubes are used for plasma collection. These samples are aliquoted and stored in 16 cryotubes (1.5 ml each) for shipping and long-term storage at -80ºC.

The procedure for stool sample collection is as follows: The patient collects the sample in a sterile feces collection container, filling it approximately halfway. The sample must be collected no more than 24 hours prior to processing and should be stored in a refrigerator at 4ºC by the patient. If the sample is collected at the hospital, it must be kept refrigerated until processing and should not be left at room temperature for more than 4 hours. Upon arrival at the hospital, three 5 ml Eppendorf tubes are labeled with the patient's information and the sample collection date. Approximately 200 mg of stool is then transferred into each of three labeled Eppendorf tubes. The tubes are placed in a plastic bag and stored at -20ºC until they are sent to the central laboratory for further analysis.

The preferred materials for tumor sampling, listed in order of priority, are as follows: A) at least 3 frozen biopsy cylinders taken at the time of metastasis diagnosis, B) FFPE (formalin-fixed, paraffin-embedded) block, C) remaining block from the metastasis diagnosis not specifically collected for the project (assumed to be FFPE), D) at least 30 slides from the metastasis block with an H&E stained section, and E) archived tumor block from the initial diagnosis.

The sample collection protocol emphasizes that it is crucial to obtain at least three cores, as some cores may contain only coagulated tissue or fat, making them unsuitable for analysis. A pathology report confirming the presence of tissue in the cylinders must be provided, with the anonymized report including the patient's study code.

For labeling, each paraffin block or slide box is properly labeled with the sample details, including the acquisition date (whether archived or new) and the patient's study code. Frozen tumor samples in cryotubes are stored at -80ºC, while FFPE blocks or slides are stored at room temperature in a plastic bag or box, respectively, until they are shipped to the central laboratory.

##### **Case Report Form**

During the first clinical visit, the oncologists participating in the study collect demographic data, clinical history, disease evaluation, current treatments, pregnancy and childbirth history, surgical history, comorbidities, and allergies from both the EHR and the patient. Bloodwork results (96 variables) are also added to the case report form (CRF).

##### At each subsequent visit, the CRF is updated with new clinical and radiological evaluations, as well as the latest bloodwork results. Anonymized CRF records are stored in a secure, ad-hoc server (ShareCRF, Spain), where oncologists can add and/or modify records through interactive forms. This system ensures that patient data is continuously updated and securely stored for analysis throughout the study.

##### Medical Imaging

At each visit, patients' tumors are examined using the imaging technique most appropriate for their clinical features (CT, MRI, PET, PET-CT, radiography, or ultrasound). The radiologist's evaluation of the results is then added to the CRF. Anonymized raw images are collected as part of the study's data and securely stored in an ad-hoc server for future analysis.

##### Hematoxylin and eosin-stained whole-slide imaging

In parallel with the processing of tumor samples, hematoxylin and eosin-stained (H&E) tumor slides are scanned at the central laboratory. Whole slides are acquired using a slide scanner (AxioScan Z1, Zeiss) with a Plan-Apochromat 20x/0.8 M27 objective, and images are captured using Zen Blue Software (V3.1 Zeiss). The whole slide imaging (WSI) data is then stored on the central laboratory’s server in czi format.

##### Smartwatch Sensor Data

The patients are instructed to use the wearable devices throughout the duration of the study to collect sensor data. These devices log various parameters, including physical activity (activity type and duration), number of steps, stress levels, heart rate, and oxygen saturation. Through the eB2 MindCare mobile application, patients can also register their emotions and pain levels at any time during the study. The raw data from the devices is securely logged to eB2 MindCare platform for further analysis.

##### Patient Reported Outcomes

A subjective evaluation of the patients' quality of life (QoL) and adverse effects is collected using the EORTC QLQ-C30^3^, GHQ28^4^ and PRO-CTCAE^5^ questionnaires. The patients' adherence to the Mediterranean diet is examined using a 14-item questionnaire^6^. These questionnaires are filled in during each clinical visit and are also continuously available to the patients through the eB2 MindCare mobile app.

Partial and global summary scores are calculated for EORTC QLQ-C30, GH28 and diet questionnaires using their corresponding manuals^4,6,7^, yielding 15, 5 and 1 features respectively.

The patients' pain levels are evaluated during each visit using a visual analogue scale (VAS), ranging from 0 to 10^8^. Patients can also add additional pain values at any time using the MindCare mobile application.

Patients’ emotions are recorded through the MindCare app, which provides a pre-defined list of 20 emotions characterized by positive, negative, or neutral valence. All the patient-generated data is securely logged to the eB2 server and is then copied locally for further analysis.

##### Tumor Genome

The biopsy samples obtained during the first visit are used for whole exome sequencing of the tumors. DNA is extracted from the FFPE samples using the DNeasy® Blood & Tissue Kit (QIAGEN, USA). DNA concentration is measured using the Qubit 1X dsDNA HS Assay Kit (ThermoFisher Scientific, USA) with the Qubit 4 Fluorometer (ThermoFisher Scientific, USA). DNA integrity is assessed using the Genomic DNA Assay Kit (Revvity, USA) on the LabChip GX Touch (Revvity, USA).

Samples with a total DNA content above 120 ng are used to prepare sequencing libraries with the KAPA HyperPlus Kit (Hoffmann-La Roche, Switzerland). The libraries are then sequenced using the NovaSeq X Plus (Illumina, USA) sequencer at a loading concentration of 150 pM in a 10B Flowcell (2x150bp). Raw fastq files generated by the sequencing service are received and stored locally.

Variant calls are generated from the raw data using the Varca pipeline^9^, which implements the GATK´s best practices^10^. Putative variants are then filtered, keeping only those passing the pipeline quality control filters, with a frequency lower than 1% in the general population (gnomAD^11^ and 1000 genomes project^12^), affecting primary transcripts and labeled as modifiers, moderate or high impact variants by Ensembl’s VEP^13^.

##### Germline Genome

A genomic profile of the patients is created using the blood samples obtained during the first medical visit. DNA is extracted from the PBMCs using the DNeasy® Blood & Tissue Kit (QIAGEN, USA). The total DNA mass is quantified using the Qubit 1X dsDNA HS Assay Kit (ThermoFisher Scientific, USA) in the Qubit 4 Fluorometer (ThermoFisher Scientific, USA). Samples with less than 250 ng of DNA are discarded, while the remaining samples are used for genotyping with the Infinium™ Global Screening Array-24 v3.0 BeadChip (Illumina, USA). Raw IDAT files generated by the genotyping service at the central laboratory are stored locally.

The IDAT files are processed using Illumina´s proprietary GenomeStudio v2.0.5 software^14^ to obtain variant call files. The genotype calls are then filtered based on the distribution of their reported quality metrics (GenotypeCall Score > 0.2 and Call Frequency > 0.9).

##### Germline epigenome

The blood samples collected during each visit are used to profile the patients’ epigenome. DNA is extracted from PBMCs using the same protocol as for genotyping. Sample quality is assessed using the same process as for the germline genome, with samples containing less than 600 ng of DNA being discarded. DNA methylation is then quantified using the EPICv2 methylation array (Illumina, USA). Raw IDAT files generated by human genotyping unit at the central laboratory are stored locally.

The raw data is processed using the SeSAME R library^15^. The intensity values are pre-processed using QCP parameters (quality-based masking probes of poor design, inference of channel for Infininum-I prbes, p-value masking using oob). Then methylation beta values are calculated for each probe.

##### Peripheral Blood Mononuclear Cells Transcriptome

Total RNA was extracted from PBMC samples using the RNeasy® Mini Kit (QIAGEN, 74104) according to the manufacturer's instructions. Sequencing libraries were prepared from 300 ng of total RNA using the QuantSeq 3' mRNA-Seq V2 Library Prep Kit (FWD) for Illumina (Lexogen, Cat.No. 191). Library preparation included reverse transcription with oligo-dT priming, followed by random-primed second strand synthesis, which incorporates 6-nt unique molecular identifiers (UMIs) for PCR duplicate removal during downstream analysis. Directional cDNA libraries, stranded in the sense orientation, were completed by PCR amplification with Unique Dual Index primers and sequenced in single-read format on an Illumina NovaSeq X platform using NovaSeq X Series Reagent Kits. The first 6 bases of read 1 (R1) corresponded to the UMI sequence. Basecalling and quality score assignment were performed using Illumina's Real Time Analysis software (RTA v2), and BCL files were converted to FASTQ format using bcl2fastq2 (Illumina). The reads were then aligned to the GRCh38 genome assembly using STAR (2.7.10b), and gene-level quantification was performed using featureCounts from the Subread package (v2.0.3). Differential gene expression analysis was conducted using pyDESeq2 (0.4.0). Genes were ranked by the Wald statistic produced by pyDESeq2 and used for pre-ranked gene set enrichment analysis with the gseapy Python library (1.1.4).

##### Gut Microbiome: metagenomics

DNA extracted from the stool samples collected at each visit throughout the study is used to profile the microbiome communities in the patients' gut. DNA extraction is performed using QIAGEN QIAamp® PowerFecal® Pro DNA Kits (QIAGEN, USA). The DNA mass per sample is determined using the Qubit 1X dsDNA HS Assay Kit (ThermoFisher Scientific, USA) in the Qubit 4 Fluorometer (ThermoFisher Scientific, USA). Samples with less than 120 ng of DNA are discarded.

DNA samples (0.5 ng) are simultaneously fragmented and tagged with sequencing adapters using the Nextera XT DNA Library Preparation Kit (Illumina, USA). Adapter-ligated libraries are completed by PCR (13 cycles) and extracted using a double-sided SPRI size selection. The libraries are then applied to an Illumina flow cell for cluster generation and sequenced on an Illumina instrument following the manufacturer’s recommendations. DNA libraries are sequenced using a NovaSeq X sequencer (Illumina, USA) with a target depth of 2 Gbases per sample. The raw fastq files generated by CNIO’s sequencing service are stored locally.

The sequencing data is processed using Kraken 2^16^ to detect and discard the host DNA and obtain microbial taxonomies. We then use Bracken^17^ to estimate the relative abundances of the detected taxa. In parallel, HumaNn is used to perform a functional profiling of the microbiome. For this, microbial reads are aligned to cluster representatives in the UniRef90 database and subsequently mapped to gene ontology (GO) terms to identify functional pathways associated with the microbial communities.

##### Blood Plasma Proteome

##### **Sample Preparation**

##### Plasma samples are diluted 1:10 with lysis buffer containing Tris-HCl (pH 8.5) and 4% SDS. To monitor digestion efficiency, four non-human proteins—Concanavalin A, GroEL, Nitroreductase, and Protein A—are added as controls. The samples are heated at 90°C for 15 minutes, cooled, and centrifuged to remove debris. The supernatant is then treated with TEAB buffer (pH 8.5) and 10 mM TCEP to reduce disulfide bonds, followed by alkylation with 20 mM CAA at room temperature in the dark to prevent disulfide bond reformation.

##### **Protein Cleanup and Digestion**

##### Automated protein cleanup and purification are performed using the SP3 beads protocol on the King Fisher Apex system. Sequential washing steps are applied to remove contaminants before enzymatic digestion. Proteins are digested at a 1:20 protein-to-enzyme ratio using LysC and Trypsin, with overnight incubation at 37°C to ensure efficient peptide generation. Post-digestion, samples are acidified with formic acid (pH < 2), and iRT peptides are added at a final concentration of 50 nmol/µL for cross-sample normalization before loading onto EvoTips.

##### **LC-MS/MS Analysis**

##### Peptides are analyzed using an Evosep One system coupled to an Orbitrap Astral mass spectrometer (Thermo Fisher Scientific). Samples are loaded onto Evotips and separated using the standard “Whisper Zoom” gradient with a throughput of 60 samples per day (SPD) on a 15 cm, 75 µm i.d. Aurora Elite TS column packed with 1.7 µm C18 beads (IonOpticks). The column temperature is maintained at 55°C. Peptides are ionized at 1.5 kV using an EASY-Spray source with a capillary temperature of 280°C. The mass spectrometer is operated in data-independent acquisition (DIA) mode, with MS1 spectra acquired at a resolution of 240,000. Precursor spectra are interspersed with a 1-second period, and ion peptides are fragmented using higher-energy collisional dissociation (HCD) at a normalized collision energy of 27. The AGC target percentages are set to 500% for both Full MS (maximum IT of 3 ms) and DIA MS/MS (maximum IT of 3 ms). A 2 m/z precursor isolation window is used with optimized window placement between 380-980 m/z. Precursor scan range is 390-980 m/z, and fragment scan range is 150-2000 m/z. Basal pooled samples spiked with the same non-human protein controls serve as quality controls to assess digestion efficiency.

##### **Data Processing and Analysis**

##### Raw files are processed using DIA-NN 1.9.2 in library-free mode against the mouse protein database (UniProtKB/Swiss-Prot, one protein per gene, 21,990 sequences). Precursor m/z range is set from 379 to 981, with default settings applied elsewhere. Protein group intensities are obtained by summing precursor quantity values from the report.tsv file, filtered with Lib.Q.Value and Lib.PG.Q.Value < 0.01. Protein group intensity data are further processed in DAPAR^18^. Normalization is performed using LOESS, and missing values are imputed using the SLSA algorithm^19^ for partially observed values and DetQuantile for values missing across entire conditions.

##### Plasma and fecal metabolome

Plasma and stool samples are shipped to General Metabolics (USA) for sample pre-processing and analysis.

**Plasma samples pre-processing**

The procedure begins by thawing the serum or plasma samples on ice for 30-60 minutes. The plasma or serum is then vortexed for approximately 15 seconds and centrifuged to remove any bubbles. A 20 µL aliquot of serum/plasma is transferred to a labeled 96-well plate, and 180 µL of 80% methanol in water is added to the sample. The plate is capped, placed on a shaker, and vigorously agitated for 15 seconds. The plate is incubated at 4°C for one hour. Following incubation, the samples are centrifuged at 4,000 rpm for 15 minutes to pellet the precipitate. The supernatant (100-400 µL) is then transferred to a shipping plate and stored at -20°C which is then shipped to General Metabolics (USA) for analysis.

**Stool samples pre-processing**

Between 30 and 80 mg from each sample are used, aiming to minimize the variation (ideally within a 2-3-fold range across all samples). The sample weight is measured by first weighing a pre-labeled 2 mL microtube, adding the frozen fecal material, and calculating the difference in mass. Homogenization is performed using a bead beater, after which the tubes are re-equilibrated at -20°C before proceeding with extraction. The extraction solvent, a mixture of 80% methanol and 20% water, is added to the homogenized fecal samples in two separate aliquots of 700 µL each. Each tube is vortexed for 15 seconds and briefly centrifuged to ensure proper mixing. After both extractions, the supernatant is pooled and transferred to labeled microtubes, and the samples are centrifuged again to remove any remaining solids. The final supernatant is collected and stored at -80°C. After completing the extraction, the samples are ready for shipping. Samples are then shipped overnight on dry ice.

**Metabolic profiling**

Metabolome profiles of the sample extracts are acquired using flow-injection mass spectrometry, following a method adapted from Fuhrer et al 2011. The instrumentation consists of an Agilent 6550 iFunnel LC-MS Q-TOF mass spectrometer in tandem with an MPS3 autosampler (Gerstel) and an Agilent 1260 Infinity II quaternary pump. The running buffer used is 60% isopropanol in water (v/v) buffered with 1 mM ammonium fluoride. Hexakis (1H, 1H, 3H-tetrafluoropropoxy)-phosphazene) (Agilent) and 3-amino-1-propanesulfonic acid (HOT) (Sigma Aldrich). The isocratic flow rate is set to 0.150 mL/min. The instrument is run in 4GHz High Resolution, negative ionization mode. Mass spectra between 50 and 1,000 m/z are collected in profile mode. 5 uL of each sample are injected twice, consecutively, within 0.96 minutes to serve as technical replicates. A pooled sample, from 92 different samples is injected periodically throughout each batch to be used as an anchor reference. Samples are acquired randomly within plates. The results received from General Metabolics consist of raw intensity data, with assigned putative metabolite identities. These results are stored locally at CNIO.

The raw intensity values are normalized by log-transforming the data and then applying a probabilistic quotient normalization (PQN): first the median value of each pooled reference sample is computed, then each metabolite intensity on each sample is divided by the corresponding median from the reference, and then each sample is divided by the median of all its quotients.

##### Blood plasma and stool samples heavy metals determination

The total contents of metals implicated in breast cancer (Pb, , W, and As)^20^ as well as elements with putative protective effects (Se, Zn, and Cu) are measured in stool and blood plasma samples using inductively coupled plasma mass spectrometry (ICP-MS) after acid digestion in a microwave oven (MSP 1000, CEM, Matheus, NC). Approximately 400 mg of blood plasma and 100 mg of lyophilized stool are weighed into Xpress vessel, and .750 mL of concentrated nitric acid (Scharlab, Spain) along with 0.250 mL of 30% (v/v) hydrogen peroxide (Dismadel, Spain) are added. The mixtures are digested at 150ºC for 20 min using a maximum power (800 W). After digestion, acidified solutions are diluted to a final volume of 5.0 mL with ultra-pure water, filtered through a 0.22 µm syringe nylon filters (Dismadel, Spain) and analyzed.

Elemental concentrations are determined using an Agilent 7700x ICP-MS instrument (Agilent Technologies, Tokyo, Japan) by the following isotopes: ^206^Pb, ^184^W, ^75^As, ^78^Se, ^66^Zn and ^63^ Cu, under continuous acquisition mode. The experimental conditions are: RF power of 1550W, sample depth of 8mm, plasma gas flow rate of 15.0 L·min^−1^, auxiliary gas flow rate of 0.90 L·min^−1^ and micromist nebulizer with a flow rate of 1.01 L·min^−1^. For Se determination, a reaction cell with H_2_ as reaction gas (4.1 L min^-1^) is used. Quantification is performed by external calibration with the isotope mentioned above.

### Internal pilot feasibility assessment and progression criteria (Go/Amend/No‑Go)

To ensure that the HDO study could be scaled to full accrual while preserving longitudinal data integrity across modalities, feasibility was operationalized a priori as an *internal pilot* embedded within the study. Feasibility was evaluated in the first 30 consecutively enrolled participants (10% of planned accrual) using a fixed database lock (data cut) and predefined operational endpoints spanning recruitment, retention, multimodal completeness, biospecimen quality, and data pipeline readiness. The feasibility framework and all thresholds were prespecified in the Statistical Analysis Plan (SAP; provided as a supplementary attachment), following published recommendations for progression criteria in internal pilot studies using a traffic‑light (RAG) approach (Green/Amber/Red) to support transparent escalation decisions.

Feasibility endpoints were grouped into five domains, each mapped to prespecified RAG thresholds (SAP, supplementary data): (1) Recruitment and representativeness, quantified as accrual rate relative to prespecified expectations and distribution across participating sites and tumor cohorts; (2) Retention/longitudinal continuity, quantified as the proportion of participants achieving prespecified minimum follow‑up milestones (e.g., completion of early scheduled timepoints within the first year) and the proportion withdrawing consent before those milestones; (3) Multimodal completeness, quantified separately for (i) *core scheduled modalities* (clinical visits/CRF fields, scheduled questionnaires, and scheduled biospecimen collections) using “due” denominators aligned with the visit schedule, and (ii) *continuous/passive digital modalities* (wearable and smartphone streams) using “observed days” denominators, where each stream’s completeness was summarized by the percentage of days meeting the prespecified “valid day” rules defined for that stream; (4) Biospecimen quality and usability, quantified as the proportion of collected samples passing modality‑specific QC gates required for downstream assays (e.g., minimum DNA yield thresholds for sequencing/arrays; proteomics/metabolomics batch QC acceptance), reported overall and by specimen type; and (5) Data pipeline readiness (latency and critical error rates), quantified as time from acquisition to availability in the central research repository (ingestion/curation latency) and the rate of critical data errors requiring re‑collection or preventing analysis.

For each feasibility endpoint, results were summarized as proportions with exact (Clopper–Pearson) 95% confidence intervals when applicable, together with complementary distributional summaries across participants (median and interquartile range for patient‑level completeness metrics, particularly for passive digital streams). “Due” events (visits, questionnaires, biospecimens) were defined prospectively by the protocol schedule; events were considered not due if a participant died or withdrew consent before the corresponding scheduled window. For continuous streams, “observed days” were defined as the days between digital onboarding (app/wearable activation) and the participant’s last evaluable day (death/withdrawal/end of follow‑up), and “valid days” were counted according to the stream‑specific validity rules prespecified in the SAP; no analytic imputation was used for feasibility endpoints beyond standard vendor signal processing intrinsic to derived wearable features.

Each endpoint was classified as Green, Amber, or Red by comparing the observed metric to its prespecified threshold(s) (SAP, supplementary data). Progression decisions were derived from the RAG profile using a rule‑based approach defined a priori: Go (scale‑up without major protocol changes) was concluded when all *critical* feasibility domains met Green thresholds and no domain was Red; Go with amendments was concluded when no critical domain was Red but one or more domains were Amber (triggering prespecified corrective actions such as enhanced onboarding, intensified troubleshooting SOPs, or scheduling/process refinements); and No‑Go (pause/redesign before further scale‑up) was concluded when any critical domain met Red thresholds or when multiple domains met Amber thresholds indicating systematic feasibility risks. Corrective actions and any procedural refinements implemented following the internal pilot were documented and version‑controlled, and subsequent accrual proceeded under the resulting decision framework.

### Privacy and data handling considerations

The project data encompasses -omic information (including genomics, transcriptomics, metagenomics, proteomics, and metabolomics), as well as physiological monitoring data, patient-reported outcomes, anonymized clinical records, laboratory test results, and medical imaging data.
The CNIO holds ultimate ownership of the data related to this project. To ensure compliance with applicable laws and regulations, CNIO has appointed a Data Protection Delegate in accordance with EU Law 2016/679, which is mandatory for institutions handling large quantities of data, particularly health-related data. Mr. Igor Pinedo, a lawyer contracted through DAC Beachcroft firm, serves as the Data Protection Delegate for CNIO, as required under Article 37.1.6 of the EU Data Protection laws. His role includes informing and advising CNIO staff regarding data-related matters, overseeing legal compliance, cooperating with authorities on data protection, and acting as a liaison between CNIO and regulatory bodies.

To ensure data security, the study includes a robust security plan aligned with the General Data Protection Regulation (GDPR) and European guidelines on data security for Big Data projects (Supplementary File 1). This security plan is focused on principles of Privacy by Design and by Default, traceability, and data portability. To further safeguard the data, external auditors have been appointed to monitor any potential data breaches or incidents that may result in data escaping CNIO’s control, while a specialized IT firm has been contracted to protect the data from cyberattacks. Any data breaches will be promptly reported to the relevant authorities and legal bodies.

The activities of the project, as well as the intended use of the data generated by the consortium, are reviewed by the Research Ethics Committees of both CNIO and the Carlos III Health Institute. EU Regulation 2016/679, which came into effect on 25 May 2018, governs the protection of individuals with regard to personal data processing and its free movement. The project adheres to the principles outlined in this regulation, ensuring full compliance with data protection laws throughout the study's lifecycle.

The study’s Data Management Plan (DMP) follows the FAIR (Findable, Accessible, Interoperable, Reusable) principles (Supplementary File 1). This DMP ensures the study complies with ethical standards and legal requirements. It ensures that the principles of data anonymization are strictly enforced, so no data can be linked back to an individual’s identity using reasonable methods. Additionally, the plan facilitates collaboration with the CNIO Biobank to establish a framework for accessing and co-governing multi-omic data. This data will be encoded and stored in standardized formats such as FASTQ, mxXML, and DICOM. The DMP also ensures that all data will be deposited in open-access repositories while safeguarding personal data protection and upholding ethical standards.

The eB2 MindCare app has the CE marking as a Class I medical device in accordance with Regulation (EU) 2017/745 (MDR).

References

1.	at <https://clinicaltrials.gov/study/NCT06590506>

2.	at <https://eb2.tech/productos/#mindcare>

3.	Aaronson, N.K. et al. *JNCI: Journal of the National Cancer Institute* **85**, 365–376 (1993).

4.	Goldberg, D.P. & Hillier, V.F. *Psychological medicine* **9**, 139–145 (1979).

5.	Basch, E. et al. *Journal of the National Cancer Institute* **106**, dju244 (2014).

6.	Martínez-González, M.A. et al. (2012).

7.	(2001).

8.	Scott, J. & Huskisson, E. *Annals of the rheumatic diseases* **38**, 560 (1979).

9.	at <https://github.com/cnio-bu/varca>

10.	der Auwera, V. *Curr. Protoc. Bioinformatics* **43**, 1 (2013).

11.	Karczewski, K.J. et al. *Nature* **581**, 434–443 (2020).

12.	Siva, N. *Nature biotechnology* **26**, 256–257 (2008).

13.	McLaren, W. et al. *Genome biology* **17**, 1–14 (2016).

14.	at <https://support.illumina.com/downloads/genomestudio-2-0.html>
